# Development and Validation of Regression-based Neuropsychological Testing Norms for Peruvian adults to detect HIV-associated Neurocognitive Impairment

**DOI:** 10.64898/2026.02.09.26345550

**Authors:** Monica M. Diaz, Kimberly Enders, Sofia Tovar-Ramirez, Yamile Rodriguez-Angeles, Valeria Roldan, Marleny Nolasco, Yating Zou, Jane She, Patricia Sotolongo, Fernando Mejia, Victor Valcour, Patricia J. Garcia, María J. Marquine, Elena Tsoy

## Abstract

**Introduction:** Neurocognitive impairment (NCI) remains common among people living with HIV (PWH), particularly in low- and middle-income countries where accurate diagnostic tools are limited. In Peru, the lack of locally validated neuropsychological (NP) normative data in Spanish poses a major barrier to diagnosing HIV-associated NCI, especially among PWH who develop NCI at younger ages. This study aimed to develop regression-based NP norms for young and middle-aged Spanish-speaking adults in Lima, Peru and validate the norms in demographically similar PWH to improve diagnostic precision of HIV-associated NCI.

**Methods:** A total of 164 healthy adults without HIV from Lima completed a comprehensive NP battery assessing memory, attention, executive function, and language, which are commonly affected in HIV-associated NCI. Multiple regression models were used to consider the influence of age, years of education, and sex on raw scores, yielding standardized demographically-adjusted norms for the population. The resulting norms were then applied to 310 PWH from Lima and then compared with previously published norms for Spanish speaking adults to evaluate performance differences.

**Results:** Age and education were the strongest predictors of performance across tests, while sex had minimal influence. Compared to people without HIV, PWH had significantly lower educational attainment (mean 12.6 vs. 13.7 years) and exhibited significantly worse performance on normed scores of Benson Figure Copy, Benson Figure Delayed Recall, Color Trails 1 and 2, Hopkins Verbal Learning Test – Revised, and WAIS-III Digit Symbol Coding, Digit Span, and Symbol Search. There were statistically significant differences between T-scores on nearly all tests between our population-specific norms and previously published norms in both directions, indicating potential over- and under-detection errors when applying norms from non-local samples.

**Discussion:** Our findings highlight the utility of locally derived norms in detecting subtle cognitive changes among young and middle-aged PWH compared with previously published norms for Spanish-speakers. Application of these norms reveals significant between-group differences that may go undetected using non-local normative data or raw scores. Future efforts should focus on rural norm development and inclusion of individuals with lower educational backgrounds in Peru and other Latin American countries.

## Introduction

Although mortality due to HIV has decreased since the introduction of antiretroviral treatment (ART), neurocognitive impairment (NCI) remains a comorbidity of importance affecting up to 50% of all people living with HIV (PWH) on ART^1^. PWH often experience impairment in concentration, memory, and executive functioning, particularly as they age, and at an earlier age than people without HIV^2–4^. With improved access to ART worldwide, HIV-associated NCI rarely progresses to dementia^1,5^. The majority of PWH reside in low- to middle-income countries (LMICs). Yet identification and management of HIV-associated NCI in LMICs remains limited due to scarcity of accurate neuropsychological (NP) tests and accompanying normative data in these regions of the world, which are critical for accurately diagnosing HIV-associated NCI^2,6^.

Certain demographic characteristics (age, sex, years of education) tend to contribute to normal variation in NP test performance not reflective of an underlying neurological condition and thus are often considered in the development of NP normative data. These demographic effects vary across cultural and language groups, and other variables that are harder to quantify (e.g., quality of education, familiarity with testing) also impact normal NP test performance. Accurate identification of neurological conditions via NP tests requires a way to estimate the impact of such factors on NP test performance. Population-specific normative data that adjust for demographic factors within a population of interest can help improve diagnostic accuracy for neurological dysfunction^7^.

HIV is a prevalent issue in Peru, a country of 34 million people with an estimated annual HIV incidence rate of 6.69%^8^. Among PWH 40 years of age or older, the prevalence of HIV-associated NCI has been reported as 28% in one cohort in Lima^9^. Yet, reliable normative data for young and middle-aged adults in Peru remains scarce, which poses a significant barrier to accurate diagnosis and care of HIV-associated NCI^10^.

While more NP tests in English have been translated to Spanish compared to other languages worldwide, normative data for Spanish-translated NP tests from a comparable healthy population are generally lacking^11^. Some studies have found that utilizing norms developed for English-speakers to interpret NP test results from native Spanish-speakers in the U.S. often result in inaccurate diagnostic conclusions on NP tests^12,13^. When comparing NP test results from native Spanish-speakers to norms validated for English speakers, NP test results fail to consider cultural, linguistic, and demographic factors that impact test performance, resulting in inaccurate diagnostic conclusions^10,12^. Similarly, despite the perception of Latinos as a relatively homogeneous group^14^, significant score differences have been observed on verbal fluency and processing speed tasks across different Latin American countries^15–17^. These factors, including regional, linguistic, socioeconomic, and cultural differences, pose significant challenges to developing homogenous NP testing norms for Spanish speakers across all of Latin America, highlighting the need for region-specific NP normative data.

Accurate interpretation of neurocognitive test performance depends on the availability of robust normative data, which provide the reference framework against which an individual’s scores are evaluated to help reduce misclassification of neurocognitive disorders (due to age, education, or other demographic effects)^18^. Although normative data have been developed for certain Peruvian populations^15,19,20^, these efforts have often been limited to specific regions, small sample sizes, or restricted age and education ranges, which may not fully capture the sociodemographic diversity of the country.

Our study builds on these prior initiatives^9^ by providing expanded, demographically adjusted normative data that account for contextual factors particularly relevant to PWH in Peru. Given PWH live longer, accurate norms allow for detection of HIV-associated NCI at earlier ages leading to earlier intervention. As the majority of PWH in Peru are adults above age 18 and below age 65, this study aimed to address a critical gap in NP normative data on a multidomain battery of cognitive tests widely used to support the diagnosis of HIV-associated NCI^2^. Specifically, we aimed to develop and test regression-based NP norms for young and middle-aged Spanish-speaking adults in Lima, Peru, to improve diagnostic precision of HIV-associated NCI in this region.

## Methods

### Study Population and Sample Selection

Participants were recruited as part of a study on cognitive and functional outcomes among adult PWH in Lima, Peru with data collected from January 2023 to March 2025. We enrolled participants living in Lima in a wide range of districts representative of the city’s population. We used stratified random sampling to ensure that our sample was representative of the broader population in terms of age, education level, and sex.

We included participants who were 18 years of age or older, native Spanish speaking, of Peruvian nationality, and having completed at least primary school (5-6 years of formal education). The sample of people without HIV was comprised of healthy men and women who tested negative using *OnSite* HIV 1/2 Antibody Plus Combo rapid antibody testing, a lateral flow immunoassay for detection of HIV-1 and 2 antibodies (IgG, IgM and IgA), at the time of their study visit and prior to beginning study procedures. The PWH sample had a history of HIV by chart review, had an undetectable HIV plasma viral load at the time of study visit, and were on ART within the prior 12 months. We excluded individuals with a history of other neurological conditions associated with NCI (i.e., stroke, epilepsy, significant traumatic brain injury, infections affecting the central nervous system), history of untreated or under-treated active psychotic illness, and self-reported lifetime (including current) substance (alcohol, marijuana, illicit substances) dependence or abuse as defined by the DSM-V^21^. A study neurologist (MMD) reviewed all participants’ eligibility criteria in which there was a question of the exclusion criteria on a case-by-case basis.

Participants without HIV were recruited through flyers posted in the healthcare setting (Hospital Cayetano Heredia) where the study took place or were family members or friends of PWH enrolled in our study. PWH were recruited from the waiting room of a public hospital HIV clinic, physician referral, or from local non-governmental organizations supporting PWH in Lima.

### Ethical Considerations

This study was approved by the University of North Carolina at Chapel Hill IRB (IRB# 22-1559), Universidad Peruana Cayetano Heredia IRB (SIDISI# 209512) and Hospital Cayetano Heredia IRB (IRB# 088-2022). All participants provided written informed consent prior to enrolling in the study.

### Study Procedures

All participants completed the NP tests in a private clinic room administered by a trained examiner in Peru. A native Spanish-speaking U.S.-licensed neuropsychologist (MJM) trained and supervised NP test administration and scoring. Quality control was performed every other month by the principal investigator (MMD), and any concerns were relayed to and resolved via consensus with the study neuropsychologist (MJM). All participants also completed standardized demographic questionnaires capturing age, sex assigned at birth, and years of education completed. All participants had been educated in Peru within the national schooling system.

### Neuropsychological Test Battery

We selected a comprehensive battery of NP tests that assess seven cognitive domains (attention/working memory, executive function, learning and memory, fine motor skills, speed of information processing, language, and visuospatial skills). The tests were chosen based on their published psychometric characteristics (validity, reliability, etc.) and using the definition of HIV-associated neurocognitive disorder by Antinori et al (2007)^2^, including several measures that were included in the AIDS Clinical Trials Group NP battery^19^. All tests were available in Spanish or had been previously translated and validated for use with Spanish-speaking populations.

The primary scores from the NP test battery included: Color Trails 2 Total Time^22^ (executive function); Wechsler Adult Intelligence Scale-III (WAIS-III) Digit Span^23^ Forward and Backward Total Score (attention/working memory); Grooved Pegboard Total Time^24^ (fine motor skills); Hopkins Verbal Learning Test – Revised^25,26^ (HVLT-R) Sum of Learning Trials and Delayed Recall Scores, Benson Complex Figure Delayed Recall Total Score^27^ (learning and memory); Benson Complex Figure Copy Total Score (visuoconstruction); Color Trails 1 Total Time^22^, WAIS-III Digit Symbol Coding Total Correct^23^, and WAIS-III Symbol Search Total Correct (processing speed); and Letter Fluency (“PRM”)^28^ and Animal Fluency^29^ Total Correct (language).

### Statistical Analyses

#### Development of Regression-based Norms among People without HIV

The NP data from participants without HIV were used to create the regression-based norms. We used a standardized regression-based approach to develop region-specific norms for Peruvian adults, as this approach has been reported to have greater accuracy of prediction and sensitivity compared to the traditional norming methods even when derived from smaller normative samples^30,31^.

Descriptive statistics were used to characterize baseline NP performance. Student’s t-tests and Wilcoxon two sample tests were used to analyze group differences by sex. Bivariate correlations were used to examine associations of NP test raw scores with age and education. To develop regression-based normative equations, we performed a set of multiple regression models with primary raw scores on individual NP tests as outcomes. The initial models included demographic predictors of age (years), age^2 (to assess non-linear effects of age), education (years), education^2 (to assess non-linear effects of education), and sex (binary, reference = female). Backward elimination was then performed to reduce the models to only significant predictors with *P-value* set at .10 to minimize Type II error. For any main effect combinations in the model that remained after backward elimination, the corresponding interaction term was added to test for potential interactions. For example, if sex and education were the only significant predictors in the model after backward elimination, then an interaction term of sex by education was tested. All final models were assessed using regression diagnostics for normality of residual distributions, homoscedasticity of the residuals, influential cases, and multicollinearity to ensure that no assumptions were violated. Based on the results from the final models, we calculated demographically adjusted z-scores for individual NP tests based on the following formula: *z = (Y – Y’)/RSE*, where *Y* is the observed raw score, *Y’* is the predicted score derived from the regression model, and *RSE* is the residual standard error of the corresponding regression equation. We then converted all z-scores to T-scores to facilitate clinical interpretation and to allow for comparison with published norms previously developed for Spanish speaking adults in the U.S.^27,32^

#### Validation of Regression-Based Norms in PWH

We then tested the performance of the regression-based norms in a demographically similar population of PWH from Lima, Peru. We applied the regression-based equations developed in the group of people without HIV to the PWH sample to characterize global cognitive performance using the HIV-associated NCI definition based on Antinori et al “Frascati” (2007) criteria defined as impairment (≥1 SD below the mean) in at least two cognitive domains^2^. We performed t-tests to compare the derived T-scores by HIV status. We also tested for differences between T-scores derived from our normative sample to those using previously published normative data for Spanish speakers using t-tests^27,32^ among PWH. All analyses were performed using STATA.

## Results

### Participant Demographic Characteristics

The total sample of this study was comprised of 474 Peruvian participants: 164 healthy adults without HIV and 310 PWH ages 18 to 80 and years of education ranging from 6 to 20 years. Mean age was similar between groups (*P*=.128), but the sample without HIV had higher educational level (*P*<.0001) and included more females (*P*=.0005) (**Table 1)**. Among 164 participants without HIV, there were no statistically significant differences in age and educational level by sex (all *P*>0.05) (**Table 2)**.

**Table 1.** Demographic characteristics and neuropsychological test T-scores in the study sample by HIV status (N=474)

|  | **HIV- (n=164)** | **PWH (n=310)** | ***P-value*** |
| --- | --- | --- | --- |
| **Demographic Characteristic (mean [SD])** |  |  |  |
| Age (years) | 45.7 (13.9) | 47.6 (11.9) | .1277 |
| Education (years) | 13.7 (2.4) | 12.6 (2.8) | **<.0001** |
| Female sex (n [%]) | 79 (48.2%) | 99 (31.9%) | **.0005** |
| **Neuropsychological Test T-scores (mean [SD])\*** |  |  |  |
| Animal Fluency | 50.0 (9.9) | 48.2 (10.3) | .0690 |
| Benson Figure Copy | 50.0 (9.9) | 44.3 (15.2) | **<.0001** |
| Benson Figure Delayed Recall | 50.0 (9.9) | 45.8 (11.6) | **<.0001** |
| Color Trails 1 | 50.0 (9.9) | 46.9 (12.5) | **.0037** |
| Color Trails 2 | 50.0 (9.9) | 45.2 (15.7) | **<.0001** |
| Letter Fluency (“PMR”) | 50.0 (9.9) | 49.9 (12.2) | .9047 |
| Grooved Pegboard (Dominant hand) | 50.0 (9.9) | 48.3 (9.6) | .0698 |
| Grooved Pegboard (Non-dominant hand) | 50.0 (9.9) | 48.3 (11.2) | .1105 |
| HVLT-R Sum of Learning Trials | 50.0 (9.9) | 47.2 (11.0) | **.0067** |
| HVLT-R Delayed Recall | 50.0 (9.9) | 46.8 (12.7) | **.0027** |
| WAIS-III Digit Symbol Coding | 50.0 (9.9) | 44.7 (10.4) | **<.0001** |
| WAIS-III Digit Span Forward and Backward | 50.0 (9.9) | 47.0 (10.2) | **.0019** |
| WAIS-III Symbol Search | 50.0 (9.9) | 46.9 (11.3) | **.0034** |

\*Applying locally derived regression-based norms derived from the group of people without HIV
**Abbreviations:** HVLT-R = Hopkins Verbal Learning Test – Revised; PWH = people living with HIV; WAIS-III = Weschler Adult Intelligence Scale-III

**Table 2.** Demographic characteristics and neuropsychological test T-scores in the reference sample of people without HIV by sex (n=164)

|  | **Females (n=79 )** | **Males (n=85 )** | ***P-value*** |
| --- | --- | --- | --- |
| **Demographic Characteristic (mean [SD])** |  |  |  |
| Age (years) | 46.0(12.3) | 45.3(15.2) | .7450 |
| Education (years) | 13.7(2.5) | 13.7(2.3) | .9631 |
| **Neuropsychological Test T-scores (mean [SD])\*** |  |  |  |
| Animal Fluency | 50.1(10.1) | 49.9(9.8) | .8804 |
| Benson Figure Copy | 50.0(10.9) | 50.0(9.1) | 1.0000 |
| Benson Figure Delayed Recall | 50.1(9.4) | 49.9(10.4) | .8736 |
| Color Trails 1 | 49.3(10.3) | 50.7(9.6) | .3637 |
| Color Trails 2 | 48.8(10.0) | 51.1(9.9) | .1371 |
| Letter Fluency (“PMR”) | 49.6(9.5) | 50.3(10.4) | .6625 |
| Grooved Pegboard (Dominant hand) | 49.0(11.2) | 50.9(8.6) | .2262 |
| Grooved Pegboard (Non-dominant hand) | 49.7(10.4) | 50.3(9.5) | .7243 |
| HVLT-R Sum of Learning Trials | 51.5(10.1) | 48.6(9.6) | .0612 |
| HVLT-R Delayed Recall | 50.2(8.8) | 49.8(11.0) | .7625 |
| WAIS-III Digit Symbol Coding | 50.7(9.9) | 49.4(10.0) | .4142 |
| WAIS-III Digit Span Forward and Backward | 50.0(9.8) | 50.0(10.1) | 1.0000 |
| WAIS-III Symbol Search | 48.6(10.3) | 51.3(9.5) | .0733 |

**Abbreviations:** HVLT-R = Hopkins Verbal Learning Test – Revised; WAIS-III = Weschler Adult Intelligence Scale-III
\*Applying locally derived regression-based norms derived from the group of people without HIV

### Normative Regression Models

**Table 3** shows the results from the final multiple linear regression model for individual NP test outcomes among healthy participants without HIV (n=164). Model R^2^ values ranged from 0.06 for Benson Complex Figure Copy to 0.43 for WAIS-III Symbol Search. Across models, age was the strongest predictor of performance, followed by educational attainment in years. A significant positive effect of male sex was found on tests of visuoconstruction (Benson Complex Figure Copy) and attention/working memory (WAIS-III Digit Span). Following application of demographic adjustments, no significant differences were found between males and females in the reference sample (**Table 2**). **Supplementary Table I** presents the full model outputs with all significant and non-significant predictors. Across most models, squared terms for age and education were not statistically significant. Squared age effects were significant for Color Trails 2 and Grooved Pegboard measures, while squared education effects were found for Benson Figure Copy and Delayed Recall and Color Trails 2. We identified statistically significant or borderline significant interactions between age and education on timed measures of Color Trails 2 (*P*=.004, *RSE*=32.1) and Grooved Pegboard (Dominant hand; *P*=.054, *RSE*=14.8). Model comparisons revealed minimal incremental increases in R^2^ values when retaining squared and interaction terms, which were subsequently dropped from final models to minimize overfitting.

**Table 3.**
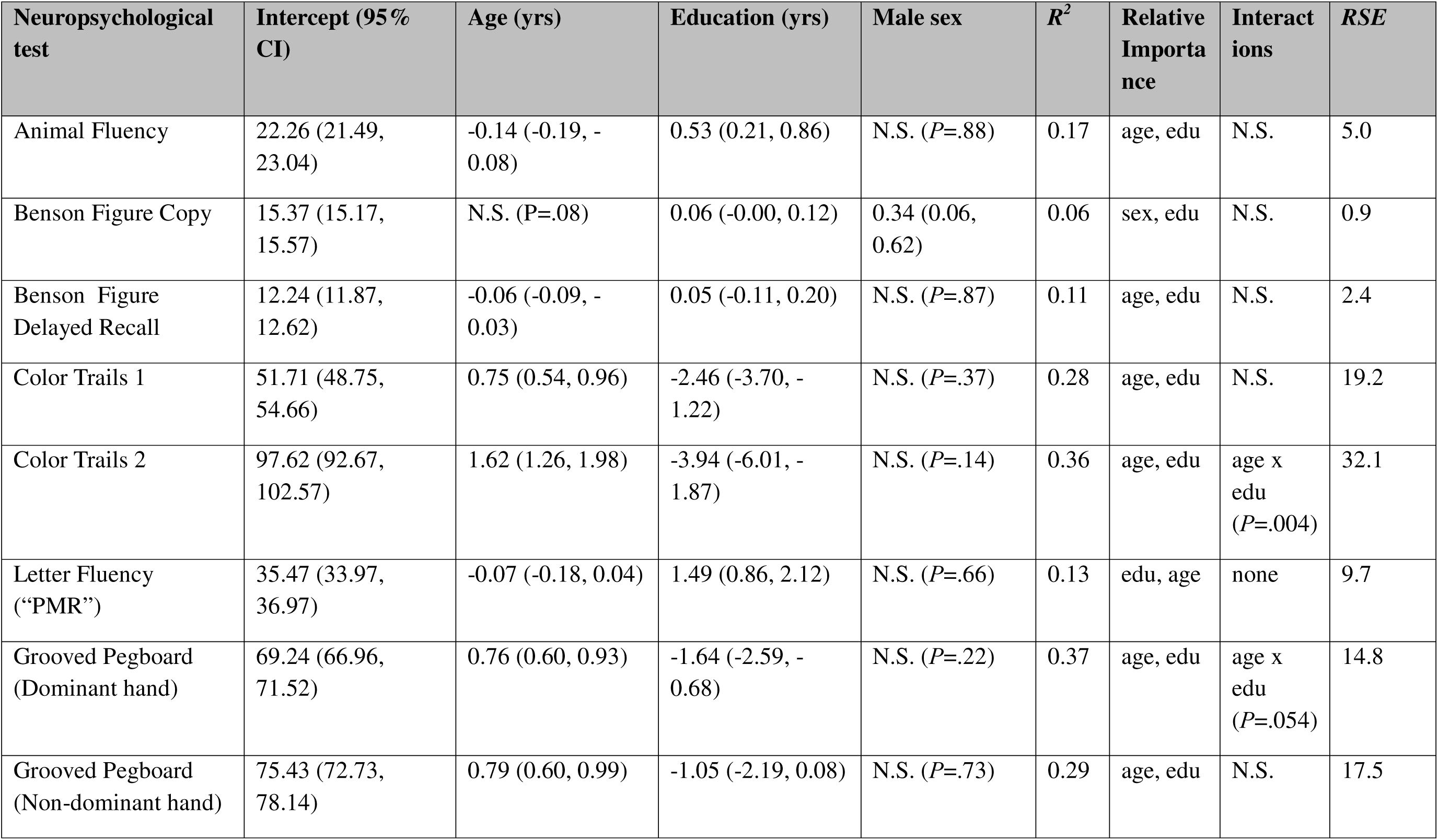
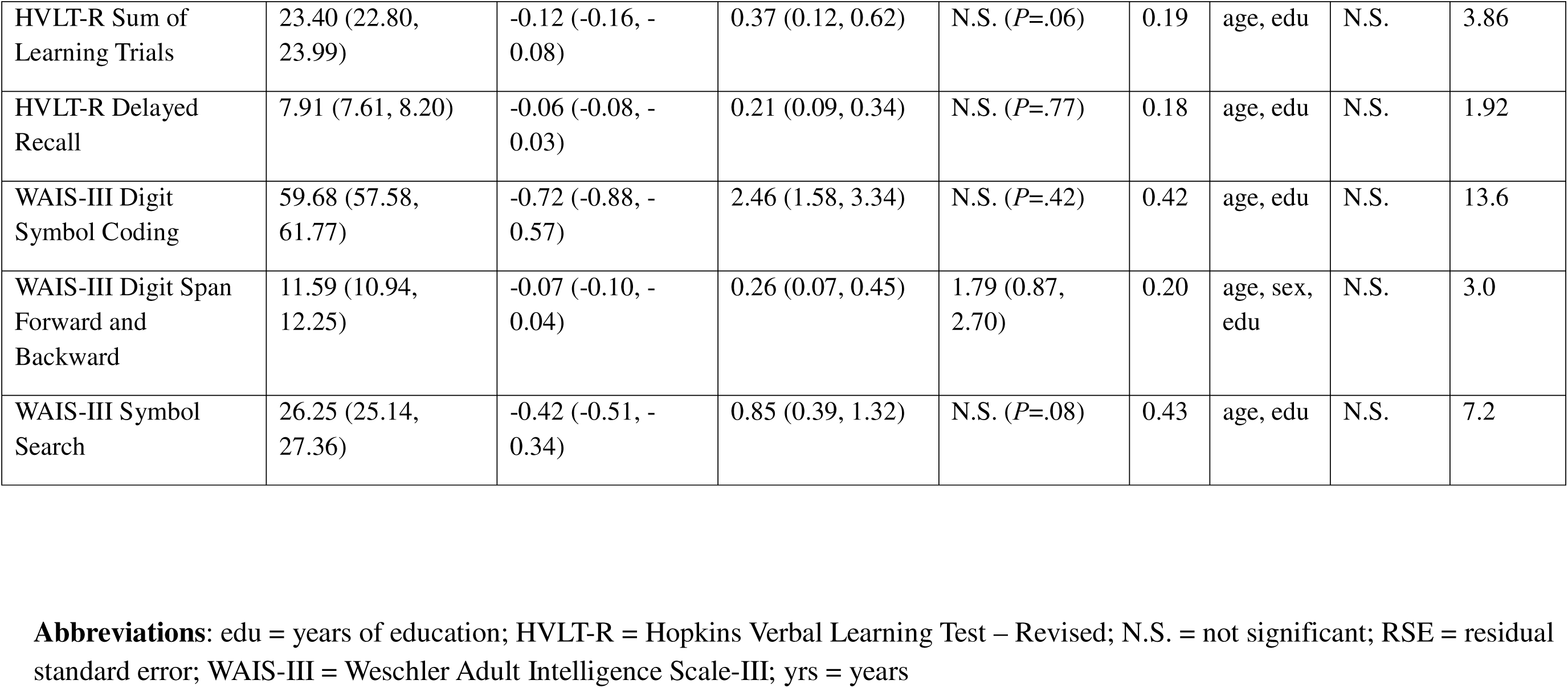
Results of linear regression models for neuropsychological test outcomes and demographic predictors in the reference sample of people without HIV (n=164)

### Demographically Adjusted Cognitive Performance by HIV Status

Demographically adjusted T-scores on individual NP tests were compared by HIV status in the entire sample **(Table 1).** On average, PWH had significantly worse performance on tests of visuoconstruction (Benson Complex Figure Copy), visual memory (Benson Complex Figure Delayed Recall), executive functions (Color Trails 2), verbal memory (HVLT-R Sum of Learning Trials and Delayed Recall), attention/working memory (WAIS-III Digit Span), and processing speed (Color Trails 1, WAIS-III Digit Symbol Coding, and WAIS-III Symbol Search). No statistically significant differences were found on verbal fluency (“PMR” and Animal Fluency) and motor (Grooved Pegboard) measures. **Supplementary Table II** demonstrates the prevalence of NP test impairment (defined as T-score < 40) in the entire sample by HIV status. These results were consistent with those including T-scores as continuous variables (worse performance among PWH on nearly all NP tests), except that more PWH were impaired on Grooved Pegboard Non-Dominant Total Time performance compared to those without HIV.

### Comparisons to previously published norms for Spanish speaking adult U.S. residents

**Table 4** demonstrates a comparison of demographically adjusted T-scores using both locally derived norms developed in this study and using previously published normative data from a sample of Spanish speaking adults in the U.S. of similar age and educational attainment. We applied these two sets of normative equations to our sample of PWH (n=310) and found statistically significant mean differences between T-scores on nearly all NP tests except Grooved Pegboard. When applying our locally derived norms, T-scores were significantly lower compared to the external sample norms on tests of visuoconstruction (Benson Figure Copy), category fluency (Animal Fluency), executive functions (Color Trails 2), attention/working memory (WAIS-III Digit Span), and one measure of processing speed (Color Trails 1). Conversely, T-scores on measures of visual and verbal memory (Benson Figure Delayed Recall, HVLT-R Sum of Learning Trials, HVLT-R Delayed Recall), letter fluency (“PMR”), and two processing speed tests (WAIS-III Digit Symbol Coding, WAIS-III Symbol Search) were significantly higher using local norms compared to previously published ones. Mean differences ranged from -3.2 (Benson Figure Delayed Recall) to 20.2 (WAIS-III Digit Span).

**Table 4.**
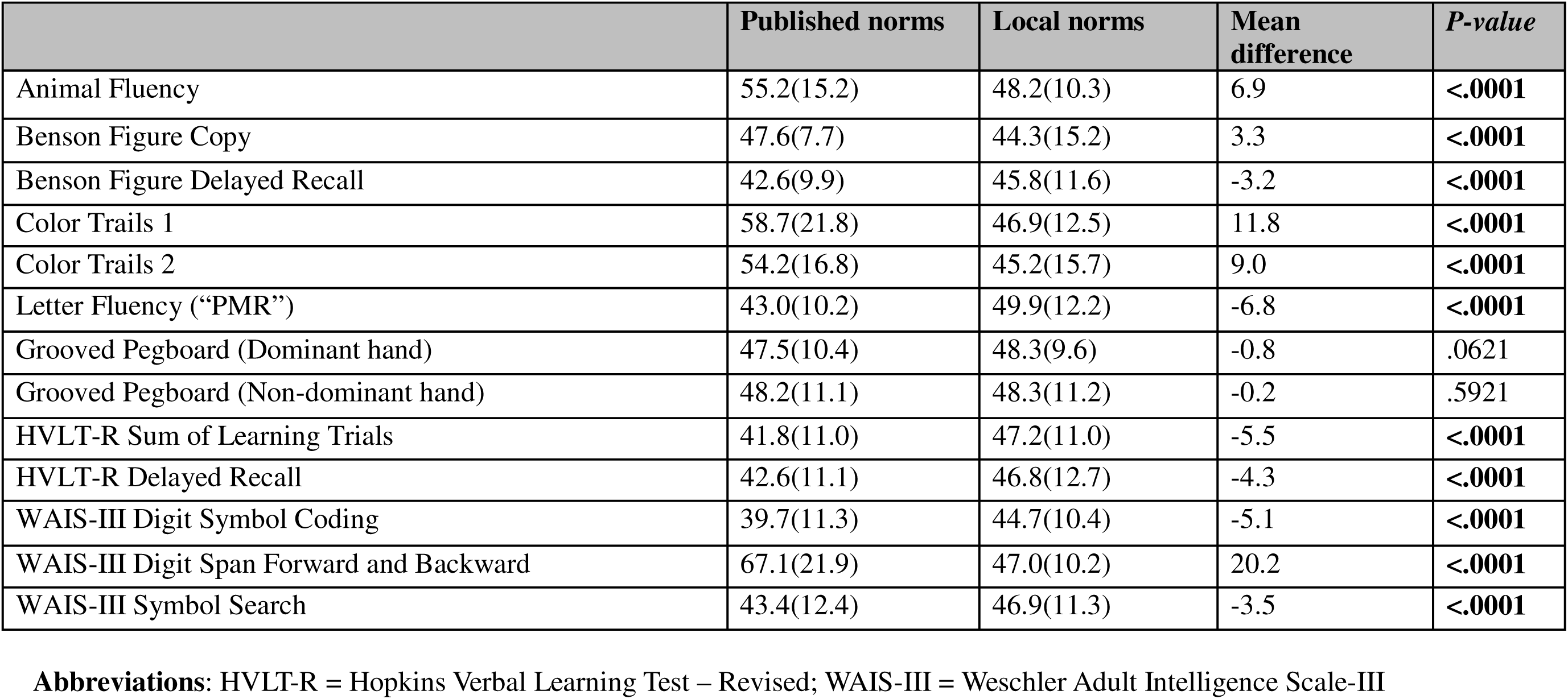
Comparison of T-scores derived from locally developed norms and published norms from an external sample of Spanish speakers in people living with HIV (n=310)

When prevalence of NP impaired scores (T-score < 40) was assessed applying the two sets of normative data among the PWH, the results were consistent with greater prevalence of impaired scores when using local norms on Animal Fluency, Benson Figure Copy, Color Trails 1 and 2, and WAIS-III Digit Span (**Supplementary Table III**). Conversely, lower rates of impaired scores using locally derived norms were seen on Benson Figure Delayed Recall, Letter Fluency “PMR,” HVLT-R Sum of Learning Trials and Delayed Recall, WAIS-III Digit Symbol Coding, WAIS-III Symbol Search, and Grooved Pegboard (both dominant and non-dominant hand conditions).

## Discussion

This study used a rigorous and systematic approach to develop normative data on a multidomain NP test battery for young and middle-aged Spanish-speaking adults in urban Peru. Consistent with prior literature, we found that performance on most tests was strongly influenced by age followed by educational level, with a minority of test results being influenced by sex. Moreover, we found that when applying these norms to a local sample of PWH, PWH performed significantly worse on a wide range of NP tests compared to HIV-negative reference participants. When we compared our locally derived norms to previously published norms from a demographically but not geographically similar sample^27,32^, we found significant discrepancies in prevalence of impaired performance in both directions. Overall, our methodology provides a robust way to generate locally derived NP test normative data that can be applied to younger and middle-aged populations in Latin America and other LMICs for adequate comparisons. This underscores the importance of developing demographically-adjusted population specific normative NP data.

In the United States, efforts are being made to include diverse populations in normative data for NP testing^12,32^. The majority of NP tests have been developed and validated in Western, industrialized populations with higher levels of education and socioeconomic status^33^. This is important to note considering that access to education and quality of education is not the same throughout LMICs^34^. Using norms developed in other countries, especially in regions with vastly different sociocultural contexts, often leads to inaccurate interpretations that fail to reflect the true cognitive abilities of the individual being tested. For example, misdiagnosis rates of cognitive impairment were up to 20% in a study testing populations in Morocco, Spain, and Colombia when norms from North American samples were used^35^. Another study found that locally developed norms more accurately classified cognitive impairment in Colombia compared with Chilean norms that had been historically used prior to the Colombian norm development^36^. Our results are consistent with these reports and highlight the potential of both over- and under-detection of cognitive impairment when applying norms from external samples, even those demographically similar and language-matched with the target population.

Although several NP normative studies have been conducted in Latin American populations, they primarily relied on data from small samples within each country and did not use rigorous regression-based norming methods^14,37,38^. One of the major challenges in generating appropriate norms for the region is the vast cultural and demographic heterogeneity of Latin American populations. Norms derived from a healthy population must accurately reflect the cognitive performance of the diverse subgroups within the population requiring sampling strategies that provide adequate representation of the patient population of interest. Our results, which are in line with existing literature showing that cognitive performance is strongly influenced by demographic factors such as education, age, and sex (on some measures), further support this premise as demographic characteristics vary widely not only between countries but also within countries across different patient populations.

Prior studies have shown that using regression-based norms with multiple predictors can yield norms using 2.5 to 5.5 times smaller samples compared with traditional “binned” norming approaches to achieve equivalent precision^39^. Regression-based norms thus offer greater statistical precision in small or resource-limited samples, which makes this approach more practical without requiring resource- and cost-prohibitive large normative datasets^40^. Regression-based norming methods have been previously applied in samples of Spanish speaking adults^38^ and children^41^ in Latin America and in Spain^42^. However, these prior studies did not use a wide range of domain-specific NP measures, especially those relevant to HIV-related NCI as our study has done.

Many studies have developed normative data for Spanish speakers, but many have not included tests often impacted by HIV. One study, for example, developed demographically corrected NP norms for South African adults using the HIV Neurobehavioral Research Center International Neurobehavioral Battery^43^. The study used data from 500 HIV-negative participants and found that applying U.S.-based norms led to a 62.2% impairment rate among healthy individuals, whereas the newly developed South African norms resulted in a 15.0% prevalence of impairment^43^. Another study focused on creating regression-based demographically corrected norms for Xhosa-speaking South Africans and similarly found that the newly developed local norms were more sensitive in detecting NCI in more cognitive domains among people with HIV compared with the U.S. test publisher norms ^44^. In Cameroon, one study established normative data for executive function and verbal fluency tests allowing for more accurate assessments of cognitive function in Cameroonian populations and facilitating better detection of HIV-associated NCI in both PWH and HIV-negative individuals^45^. Moreover, an AIDS Clinical Trials Group study (the International Neurocognitive Normative Study) was conducted to address the lack of neurocognitive normative data in LMICs with participants enrolled across seven countries, including a subset from Peru, using non-regression based methodology. However, the published normative data from this study was merged across countries and not available individually for each country^19^. Our study fills these gaps and provides normative data for young to middle-aged adults from Lima, Peru, using an NP battery tailored to detect HIV-associated NCI that has been lacking.

Our study has several limitations. Firstly, the sample of healthy control participants was a small sample with a broad age range and a mean educational level of 13.7 years with very few individuals with less than 10 years of education. Moreover, since we intentionally enrolled healthy control participants demographically matched to the PWH who tend to be younger, the normative sample included very few individuals over age 70. Thus, our results should be applied and interpreted with caution in people who are either over age 70 or have less than 10 years of education and should not be used for persons with less than a primary education (5-6 years) who were not included in the sample. Next, our norms were created in a sample of participants recruited via different methods from the capital city of Lima, the population of which tends to have a higher socioeconomic status compared with rural regions, limiting generalizability to non-urban samples. Our study may be replicated in other Latin American cities to create local regression-based norms for the country. At the same time, a major strength of our study is that our norms address a substantial gap in NP normative data for young and middle-aged adults, as most existing norms in Latin America were developed for pediatric or older adult populations.

## Conclusions

We developed robust normative data for young and middle-aged adults in Lima, Peru, that can facilitate accurate detection of HIV-associated NCI, which tends to have an onset at younger ages compared with more common neurodegenerative disorders of older age. Further studies are needed to refine and advance these norms in rural populations, non-Spanish speaking Peruvians, and adults with lower educational attainment and literacy levels and to develop norms for other regions throughout Latin America.

## Data Availability

Data and materials can be made available by request to the corresponding author.

## Declarations

## Ethics approval and consent to participate

In accordance with the Declaration of Helsinki, this study was approved by the University of North Carolina at Chapel Hill IRB (IRB# 22-1559), Universidad Peruana Cayetano Heredia IRB (SIDISI# 209512) and Hospital Cayetano Heredia IRB (IRB# 088-2022). All participants provided written informed consent prior to enrolling in the study.

## Consent for publication

Not applicable

## Availability of data and materials

Data and materials can be made available by request to the corresponding author.

## Authors’ contributions

MMD- drafting or original manuscript; overall supervision and guidance of the study; review of final draft of manuscript

KE- data analysis; review of final draft of manuscript

ST- data collection; review of final draft of manuscript

YR- data collection; review of final draft of manuscript

VR- data collection; review of final draft of manuscript

MN- data collection; review of final draft of manuscript

YZ- data analysis; review of final draft of manuscript

JS- data analysis; review of final draft of manuscript

PS- overall supervision and guidance of the study; review of final draft of manuscript

FM- overall supervision and guidance of the study; review of final draft of manuscript

VV- overall supervision and guidance of the study; review of final draft of manuscript

PJG- overall supervision and guidance of the study; review of final draft of manuscript

MJM- overall supervision and guidance of the study; review of final draft of manuscript

ET- overall supervision and guidance of the study; review of final draft of manuscript

## Acknowledgements

We would like to thank the participants who participated in this study.

## References

1. Heaton, R. K. et al. HIV-associated neurocognitive disorders before and during the era of combination antiretroviral therapy: differences in rates, nature, and predictors. J Neurovirol 17, 3–16 (2011).

2. Antinori, A. et al. Updated research nosology for HIV-associated neurocognitive disorders. Neurology 69, 1789–1799 (2007).

3. Brouillette, M.-J. et al. Identifying Neurocognitive Decline at 36 Months among HIV-Positive Participants in the CHARTER Cohort Using Group-Based Trajectory Analysis. PLoS One 11, e0155766 (2016).

4. Aung, H. L. et al. Cognitive ageing is premature among a community sample of optimally treated people living with HIV. HIV Med 22, 151–164 (2021).

5. Lam, J. O. et al. Comparison of dementia risk after age 50 between individuals with and without HIV infection. AIDS 35, 821–828 (2021).

6. Nightingale, S. et al. Cognitive impairment in people living with HIV: consensus recommendations for a new approach. Nat Rev Neurol 19, 424–433 (2023).

7. Marquine, M. J. et al. Introduction to the Neuropsychological Norms for the US-Mexico Border Region in Spanish (NP-NUMBRS) Project. Clin Neuropsychol 35, 227–235 (2021).

8. Torres, T. S. et al. Recent HIV infection and annualized HIV incidence rates among sexual and gender minorities in Brazil and Peru (ImPrEP seroincidence study): a cross-sectional, multicenter study. Lancet Reg Health Am 28, 100642 (2023).

9. Diaz, M. M. et al. Characterization of HIV-Associated Neurocognitive Impairment in Middle-Aged and Older Persons With HIV in Lima, Peru. Frontiers in Neurology 12, 795 (2021).

10. Morlett Paredes, A., et al. The state of neuropsychological test norms for Spanish-speaking adults in the United States. Clin Neuropsychol 35, 236–252 (2021).

11. Strutt, A. M. et al. Culturally and Linguistically Informed Neuropsychological Evaluation Protocol for Primarily Spanish-Speaking Adults. Arch Clin Neuropsychol 38, 408–432 (2023).

12. Morlett Paredes, A., et al. Demographically adjusted normative data for the Halstead category test in a Spanish-speaking adult population: Results from the Neuropsychological Norms for the U.S.-Mexico Border Region in Spanish (NP-NUMBRS). Clin Neuropsychol 35, 356–373 (2021).

13. Casaletto, K. B. et al. Demographically Corrected Normative Standards for the Spanish Language Version of the NIH Toolbox Cognition Battery. J Int Neuropsychol Soc 22, 364–374 (2016).

14. Pontón, M. O. & Ardila, A. The future of neuropsychology with Hispanic populations in the United States. Arch Clin Neuropsychol 14, 565–580 (1999).

15. Rivera, D. et al. Normative data for verbal fluency in healthy Latin American adults: Letter M, and fruits and occupations categories. Neuropsychology 33, 287–300 (2019).

16. Arango-Lasprilla, J. C. et al. Symbol Digit Modalities Test: Normative data for the Latin American Spanish speaking adult population. NeuroRehabilitation 37, 625–638 (2015).

17. Rivera, D. et al. Brief Test of Attention: Normative data for the Latin American Spanish speaking adult population. NeuroRehabilitation 37, 663–676 (2015).

18. Casaletto, K. B. et al. Demographically Corrected Normative Standards for the English Version of the NIH Toolbox Cognition Battery. J Int Neuropsychol Soc 21, 378–391 (2015).

19. Robertson, K. et al. International neurocognitive normative study: neurocognitive comparison data in diverse resource-limited settings: AIDS Clinical Trials Group A5271. J Neurovirol 22, 472–478 (2016).

20. Bulgarelli, L. et al. Normative data of the Spanish version of the Montreal Cognitive Assessment (MoCA) in older individuals from Peru. Dement Neuropsychol 19, e20240261 (2025).

21. Hasin, D. S. et al. DSM-5 criteria for substance use disorders: recommendations and rationale. Am J Psychiatry 170, 834–851 (2013).

22. García-Escobar, G. et al. Spanish normative studies (NEURONORMA-Plus project): norms for the Delis Kaplan-Design Fluency Test, Color Trails Test, and Dual Task. Neurologia (Engl Ed) 39, 160–169 (2024).

23. Wechsler, D. (2001). WAIS-III Escala de Inteligencia de Wechsler para Adultos-III: Manual técnico (2a ed., adaptación española: Dpto. I+D, TEA Ediciones, S.A.). Madrid: TEA Ediciones.

24. Lafayette Instrument 32025 Manual de usuario de prueba de tablero perforado ranurado Lafayette Instrument 32025 Grooved Pegboard Test User Manual. https://manuals.plus/es/lafayette-instrument/32025-grooved-pegboard-test-manual.

25. Brandt J, Benedict RHB. Hopkins Verbal Learning Test -- Revised Professional Manual. Lutz, FL: Psychological Assessment Resources, Inc; 2001.

26. González-Palau, F. et al. Clinical utility of the hopkins Verbal Test-Revised for detecting Alzheimer’s disease and mild cognitive impairment in Spanish population. Arch Clin Neuropsychol 28, 245–253 (2013).

27. Marquine, M. J. et al. Demographically-adjusted normative data among Latinos for the version 3 of the Alzheimer’s Disease Centers’ Neuropsychological Test Battery in the Uniform Data Set. Alzheimers Dement 19, 4174–4186 (2023).

28. Sáez-Atxukarro, O. et al. [Hopkins Verbal Learning Test-revised: normalization and standardization for Spanish population]. Rev Neurol 72, 35–42 (2021).

29. Benton A, Hamsher K, Sivan A. Multilingual Aphasia Examination (3rd edition). Iowa City, IA: AJA Associates; 1983.

30. Van der Elst, W., Hurks, P., Wassenberg, R., Meijs, C. & Jolles, J. Animal Verbal Fluency and Design Fluency in school-aged children: effects of age, sex, and mean level of parental education, and regression-based normative data. J Clin Exp Neuropsychol 33, 1005–1015 (2011).

31. Shirk, S. D. et al. A web-based normative calculator for the uniform data set (UDS) neuropsychological test battery. Alzheimers Res Ther 3, 32 (2011).

32. Cherner, M. et al. Neuropsychological Norms for the U.S.-Mexico Border Region in Spanish (NP-NUMBRS) Project: Methodology and sample characteristics. Clin Neuropsychol 35, 253–268 (2021).

33. Fernández, A. L. et al. The multicultural neuropsychological scale (MUNS): validity, reliability, normative data and cross-cultural evidence. Culture and Brain 10, 167–193 (2022).

34. Hossain, N. & Hickey, S. 1The Problem of Education Quality in Developing Countries. in The Politics of Education in Developing Countries: From Schooling to Learning (eds Hickey, S. & Hossain, N.) 0 (Oxford University Press, 2019). doi:10.1093/oso/9780198835684.003.0001.

35. Daugherty, J. C., Puente, A. E., Fasfous, A. F., Hidalgo-Ruzzante, N. & Pérez-Garcia, M. Diagnostic mistakes of culturally diverse individuals when using North American neuropsychological tests. Appl Neuropsychol Adult 24, 16–22 (2017).

36. Duggan, E. C., Awakon, L. M., Loaiza, C. C. & Garcia-Barrera, M. A. Contributing Towards a Cultural Neuropsychology Assessment Decision-Making Framework: Comparison of WAIS-IV Norms from Colombia, Chile, Mexico, Spain, United States, and Canada. Arch Clin Neuropsychol 34, 657–681 (2019).

37. Arango-Lasprilla, J. C. Commonly used neuropsychological tests for Spanish speakers: Normative data from Latin America. NeuroRehabilitation 37, 489–491 (2015).

38. Guàrdia-Olmos, J., Peró-Cebollero, M., Rivera, D. & Arango-Lasprilla, J. C. Methodology for the development of normative data for ten Spanish-language neuropsychological tests in eleven Latin American countries. NeuroRehabilitation 37, 493–499 (2015).

39. Oosterhuis, H. E. M., van der Ark, L. A. & Sijtsma, K. Sample Size Requirements for Traditional and Regression-Based Norms. Assessment 23, 191–202 (2016).

40. Oosterhuis, H. E. M., van der Ark, L. A. & Sijtsma, K. Sample Size Requirements for Traditional and Regression-Based Norms. Assessment 23, 191–202 (2016).

41. Rivera, D. et al. Regression-Based Normative Data for Children From Latin America: Phonological Verbal Fluency Letters M, R, and P. Assessment 28, 264–276 (2021).

42. Iñesta, C., Oltra-Cucarella, J. & Sitges-Maciá, E. Regression-Based Normative Data for Independent and Cognitively Active Spanish Older Adults: Verbal Fluency Tests and Boston Naming Test. Int J Environ Res Public Health 19, (2022).

43. Deist, M. et al. Neuropsychological Test Norms for the Assessment of HIV-Associated Neurocognitive Impairment Among South African Adults. AIDS Behav 27, 3080–3097 (2023).

44. Gouse, H. et al. Generating fair, reliable, and accurate neuropsychological test norms for people with HIV in a low- or middle-income country. J Neurovirol https://doi.org/10.1007/s13365-024-01235-6 (2024) doi:10.1007/s13365-024-01235-6.

45. Kanmogne, G. D. et al. Attention/Working Memory, Learning and Memory in Adult Cameroonians: Normative Data, Effects of HIV Infection and Viral Genotype. J Int Neuropsychol Soc 26, 607–623 (2020).

