## Supplemental Tables for "Development and Validation of Regression-based Neuropsychological Testing Norms for Peruvian adults to detect HIV-associated Neurocognitive Impairment"

**Supplementary Table I. Full output of linear regression models for neuropsychological test outcomes and demographic predictors in the reference sample of people without HIV (n=164)**

| **Neuropsychological test** | **Intercept** | **Age (yrs)** | **Age^2^** | **Education (yrs)** | **Education^2^** | **Male sex** | **R^2^** | **Relative Importance** | **Interactions** |
| --- | --- | --- | --- | --- | --- | --- | --- | --- | --- |
| Animal Fluency | 22.26 (21.49, 23.04) | -0.14 (-0.19, -0.08) | N.S. (P=0.11) | 0.53 (0.21, 0.86) | N.S. (P=0.41) | N.S. (P=0.90) | 0.17 | age, edu | N.S. |
| Benson Figure Copy | 15.49 (15.26, 15.72) | N.S. (P=0.17) | N.S. (P=0.07) | N.S. (P=0.09) | -0.02 (-0.04, -0.00) | 0.32 (0.04, 0.60) | 0.06 | sex, edu_sq | N.S. |
| Benson Figure Delayed Recall | 12.46 (12.00, 12.92) | -0.05 (-0.08, -0.03) | N.S. (P=0.44) | N.S. (P=0.20) | -0.04 (-0.09, 0.01) | N.S. (P=0.95) | 0.12 | age, edu_sq | N.S. |
| Color Trails 1 | 51.71 (48.75, 54.66) | 0.75 (0.54, 0.96) | N.S. (P=0.39) | -2.46 (-3.70, -1.22) | N.S. (P=0.10) | N.S. (P=0.33) | 0.28 | age, edu | N.S. |
| Color Trails 2 | 91.38 (83.16, 99.59) | 1.46 (1.11, 1.82) | 0.04 (0.01, 0.06) | -2.60 (-4.67, -0.52) | 0.78 (0.12, 1.43) | -9.67 (-19.29, -0.05) | 0.44 | age, age_sq, edu, edu_sq, sex | age x edu (P= .004) |
| Letter Fluency (“PMR”) | 35.47 (33.97, 36.97) | -0.07 (-0.18, 0.04) | N.S. (P=0.46) | 1.49 (0.86, 2.12) | N.S. (P=0.30) | N.S. (P=0.53) | 0.13 | edu, age | N.S. |
| Grooved Pegboard (Dominant hand) | 66.17 (63.14, 69.20) | 0.75 (0.59, 0.91) | 0.02 (0.01, 0.03) | -1.37 (-2.32, -0.42) | N.S. (P=?0.33) | N.S. (P=?0.08) | 0.4 | age, age_sq, edu | age x edu (P=.054) |
| Grooved Pegboard (Non-dominant hand) | 71.14 (67.59, 74.68) | 0.77 (0.58, 0.96) | 0.02 (0.01, 0.03) | N.S. (P=0.19) | N.S. (P=0.59) | N.S. (P=0.29) | 0.33 | age, age_sq | N.S. |
| HVLT-R Sum of Learning Trials | 23.40 (22.80, 23.99) | -0.12 (-0.16, -0.08) | N.S. (P=0.66) | 0.37 (0.12, 0.62) | N.S. (P=0.83) | N.S. (P=0.08) | 0.19 | age, edu | N.S. |
| HVLT-R Delayed Recall | 7.91 (7.61, 8.20) | -0.06 (-0.08, -0.03) | N.S. (P=?0.92) | 0.21 (0.09, 0.34) | N.S. (P=0.47) | N.S. (P=?0.79) | 0.18 | age, edu | N.S. |
| WAIS-III Digit Symbol Coding | 59.68 (57.58, 61.77) | -0.72 (-0.88, -0.57) | N.S. (P=0.65) | 2.46 (1.58, 3.34) | N.S. (P=0.61) | N.S. (P=0.50) | 0.42 | age, edu | N.S. |
| WAIS-III Digit Span Forward and Backward | 11.59 (10.94, 12.25) | -0.07 (-0.10, -0.04) | N.S. (P=0.75) | 0.26 (0.07, 0.45) | N.S. (P=0.92) | 1.79 (0.87, 2.70) | 0.2 | age, sex, edu | N.S. |
| WAIS-III Symbol Search | 26.25 (25.14, 27.36) | -0.42 (-0.51, -0.34) | N.S. (P=0.55) | 0.85 (0.39, 1.32) | N.S. (P=0.62) | N.S. (P=0.07) | 0.43 | age, edu | N.S. |

**Abbreviations**: edu = years of education; HVLT-R = Hopkins Verbal Learning Test – Revised; N.S. = not significant; RSE = residual standard error; WAIS-III = Weschler Adult Intelligence Scale-III; yrs = years

**Supplementary Table II. Prevalence of neuropsychological test impairment (T-score < 40) in the study sample by HIV status (N=474)**

|  | **HIV- (n=164)** | **PWH (n=310)** | ***P-value*** |
| --- | --- | --- | --- |
| **NP test impairment rate (n [%])*** | | | |
| Animal Fluency | 25(5.3%) | 53(11.2%) | .6048 |
| Benson Figure Copy | 20(4.2%) | 83(17.5%) | **.0002** |
| Benson Figure Delayed Recall | 32(6.8%) | 91(19.2%) | **.0201** |
| Color Trails 1 | 15(3.2%) | 73(15.4%) | **.0001** |
| Color Trails 2 | 22(4.6%) | 81(17.1%) | **.0014** |
| Letter Fluency (“PMR”) | 27(5.7%) | 59(12.4%) | .4900 |
| Grooved Pegboard (Dominant hand) | 22(4.7%) | 52(11.0%) | .3307 |
| Grooved Pegboard (Non-dominant hand) | 13(2.7%) | 45(9.5%) | **.0362** |
| HVLT-R Sum of Learning Trials | 25(5.3%) | 80(16.9%) | **.0084** |
| HVLT-R Delayed Recall | 25(5.3%) | 91(19.2%) | **.0006** |
| WAIS-III Digit Symbol Coding | 20(4.2%) | 98(20.7%) | **<.0001** |
| WAIS-III Digit Span Forward and Backward | 26(5.5%) | 82(17.3%) | **.0084** |
| WAIS-III Symbol Search | 26(5.5%) | 75(15.8%) | **.0349** |

*****Applying locally derived regression-based norms derived from the group of people without HIV

**Abbreviations**: HVLT-R = Hopkins Verbal Learning Test – Revised; PWH = people living with HIV; WAIS-III = Weschler Adult Intelligence Scale-III

**Supplementary Table III. Comparison of the prevalence rates of neuropsychological test impairment (T-score < 40) using locally developed norms and published norms from an external sample of Spanish speakers in people living with HIV (n=310)**

|  | **Published norms** | **Local norms** | ***P-value*** |
| --- | --- | --- | --- |
| Animal Fluency | 39(12.8%) | 53(17.1%) | **.0016** |
| Benson Figure Copy | 38(12.3%) | 83(26.9%) | **<.0001** |
| Benson Figure Delayed Recall | 110(35.6%) | 91(29.4%) | **<.0001** |
| Color Trails 1 | 28(9.1%) | 73(23.5%) | **<.0001** |
| Color Trails 2 | 48(15.5%) | 81(26.1%) | **.0037** |
| Letter Fluency (“PMR”) | 113(37.0%) | 59(19.0%) | **<.0001** |
| Grooved Pegboard (Dominant hand) | 70(23.1%) | 52(16.8%) | **.0080** |
| Grooved Pegboard (Non-dominant hand) | 66(22.1%) | 45(14.6%) | **<.0001** |
| HVLT-R Sum of Learning Trials | 122(40.4%) | 80(25.8%) | **<.0001** |
| HVLT-R Delayed Recall | 134(44.5%) | 91(29.4%) | **<.0001** |
| WAIS-III Digit Symbol Coding | 160(53.5%) | 98(31.6%) | **<.0001** |
| WAIS-III Digit Span Forward and Backward | 21(6.8%) | 82(26.5%) | **<.0001** |
| WAIS-III Symbol Search | 120(40.1%) | 75(24.2%) | **<.0001** |

**Abbreviations**: HVLT-R = Hopkins Verbal Learning Test – Revised; WAIS-III = Weschler Adult Intelligence Scale-III
